# State-level COVID-19 Trend Forecasting Using Mobility and Policy Data

**DOI:** 10.1101/2021.01.04.21249218

**Authors:** Yifei Wang, Hao (Michael) Peng, Long Sha, Zheyuan Liu, Pengyu Hong

## Abstract

The importance of pandemic forecast cannot be overemphasized. We propose an interpretable machine learning approach for forecasting pandemic transmission rates by utilizing local mobility statistics and government policies. A calibration step is introduced to deal with time-varying relationships between transmission rates and predictors. Experimental results demonstrate that our approach is able to make accurate two-week ahead predictions of the state-level COVID-19 infection trends in the US. Moreover, the models trained by our approach offer insights into the spread of COVID-19, such as the association between the baseline transmission rate and the state-level demographics, the effectiveness of local policies in reducing COVID-19 infections, and so on. This work provides a good understanding of COVID-19 evolution with respect to state-level characteristics and can potentially inform local policymakers in devising customized response strategies.

## 1 Introduction

The novel coronavirus disease 2019 (COVID-19) has caused a global pandemic (Zhu et al., 2020) and imposes unprecedented challenges on governments and societies around the world. The COVID-19 outbreak has two key features: high covertness and high transmissibility (Hao et al., 2020), which have pushed some healthcare systems to the brink of collapse and have prompted governments to impose strict policies on physical isolation and travel restrictions so as to mitigate the spread of COVID-19. It is hence essential to accurately predict the spread of COVID-19. Such research, especially research on local trend forecasting, can provide valuable insights to help local authorities prepare their health systems and deploy appropriate policies to mitigate the spread.

Conventional mathematical modelling of infectious diseases in epidemiology, such as compartmental models (Kermack & McKendrick, 1927) and their derivatives (Hao et al., 2020; Croccolo & Roman, 2020; Palladino et al., 2020), have been used to reconstruct and forecast transmission dynamics at macro levels. Such a model usually builds an ODE system in a top-down way to approximate the epidemic process and estimate model parameters via Monte Carlo methods. Accurate domain knowledge is required to design appropriate compartments as well as their relationships.

It is highly desired to develop models that not only capture dynamic relationships between infectious data and population but also make accurate forecasts on the spread of the disease. Oliver et al. (2020) argued that human behavior, especially mobility and physical co-presence, was necessary for spread analysis during all stages of a pandemic life cycle. Thanks to the pervasive mobile devices, in-time mobility statistics can now be obtained at a large scale. In fact, mobility information had been successfully used in building epidemiological models for H1N1 flu outbreaks (Balcan et al., 2009). This work estimates the reproduction number based on high-quality population mobility patterns, so as to make up missing incidence data during the early phase of the H1N1 pandemic. It was able to uncover the seasonal transmission potential of H1N1 in affected countries at the early stage, using mobility and transportation data worldwide in addition to the raw count of cases.

### Present work

In this work, we tackle the problem of forecasting the state-level daily new cases of COVID-19 in the US. We have developed a machine learning approach to estimate the state-level daily transmission rates via robust regression on local mobility statistics and government policies. The predicted daily transmission rates can then be accumulated to estimate the daily new cases. There are temporal variances in population behaviors (e.g., awareness of conditions relating to public health, compliance to policies, etc.), which, if not considered, can greatly affect the performance of our approach. To deal with this problem, we added a novel calibration step to our modeling, which assumes the relationships between the transmission rate variable and its predictors remain unchanged within a short time window. Empirical studies show that our approach can make satisfying predictions two weeks into the future for most states. Furthermore, our approach is well interpretable and offers insights into the spread of this pandemic. For example, we show that the baseline transmission rate, which is indicated as the bias terms in our trained models, is highly associated with state-level demographics. In addition, the factors identified to be significant in making predictions are quite consistent across states with how people and governments fight against COVID-19.

## 2 Related work

Previous works on COVID-19 trend prediction can be roughly categorized into the following two types.

### Compartmental models

Most recent approaches for COVID-19 spread analysis in epidemiology are derivatives of the *Susceptible Infectious Recovered* (SIR) model (Kermack & McKendrick, 1927; Harko et al., 2014), a widely used compartmental model. These approaches group the subjects in the system of interest into different population compartments when modeling epidemic spread. The dynamics of the system is characterized by the transitions of subjects between compartments, which are mathematically expressed as a set of differential equations. Croccolo & Roman (2020) extended the SIR model to encompass the effects of lockdown policy and applies it to COVID-19 in the US. Palladino et al. (2020) improved the standard SIR model to have a varying diffusion velocity of virus, which accounts for nonpharmaceutical interventions, and applied the model to COVID-19 in Italy. In another work, Hao et al. (2020) proposed a SAPHIRE model that contains seven compartments (susceptible, exposed, presymptomatic infectious, ascertained infectious, unascertained infectious, isolation in hospital and removed) to reconstruct transmission dynamics of COVID-19 in Wuhan, China between 01/01/2020 and 08/03/2020. This time period was divided into 5 segments (Pan et al., 2020), in each of which, the ascertainment rate and transmission rate were assumed to be fixed. They also assumed a constant population size and a constant number of travellers in each period. Fernaández-Villaverde & Jones (2020) incorporated social distancing into a SIRD (SusceptibleInfectious-Recovered-Dead) model that allows a time-varying contact rate so as to capture changes associated with social distancing and quarantine policy. They conducted simulations of deaths on various regions, such as New York City, Italy, Sweden and Spain. Picchiotti et al. (2020) built a SEIR (Susceptible-Exposed-Infected-Recovered) model that considers both personal protective measures and mobility restrictions represented as decreasing logistic functions. Chang et al. (2020) introduced a metapopulation SEIR model that integrated fine-grained, dynamic mobility networks to simulate the spread of SARS-CoV-2 in 10 of the largest US metropolitan statistical areas. The IHME COVID-19 Forecasting Team (2020) proposed a deterministic SEIR framework to model possible trajectories of COVID-19 infections and the effects of non-pharmaceutical interventions in the US at the state level. Dandekar & Barbastathis (2020) augmented the SIR model to include a time varying quarantine strength term, which is learned by a neural network from real data. Yang et al. (2020b) compared a set of SIR based models (e.g., SIR, SEIR, SEIR-AHQ (Tang et al., 2020), SEIR-QD (Peng et al., 2020), SEIR-PO (proposed), etc.) on their forecast abilities using the daily reported confirmed infected case data from the China CDC. It is evident that most compartmental models focus on dynamics reconstruction and lack the ability to make long-term predictions.

### Machine learning models

Machine learning approaches are very capable of learning complex dynamic patterns and relationships directly from data. Punn et al. (2020) and Tuli et al. (2020) applied machine learning techniques (e.g., support vector regression, polynomial regression, robust Weibull fitting, etc.) to fit the nationwide epidemic curve without considering any exogenous factors. Yang et al. (2020a) proposed a GRU-based framework for state-level trend prediction, integrating the time-varying epidemic information with environmental factors. This work incorporated static external factors including local population and age structure while ignoring dynamic population behaviors. Kapoor et al. (2020) developed a GNN-based approach for county-level COVID-19 forecasting, where Google’s human mobility data across all counties in the US are represented as a single large-scale spatio-temporal graph. Nevertheless, it did not consider other significant external factors (e.g., mandatory or voluntary mask policies). Ramchandani et al. (2020) divided county-level weekly rises of confirmed COVID-19 cases into 4 coarse categories and developed DeepCOVIDNet based on the DeepFM (Guo et al., 2017) framework that used the demographic statistics and cross-county mobility data provided by SafeGraph to make coarse-level predictions.

## 3 Results

We applied our approach to predict the state-level infection trends of COVID-19 in the US. The results reveal the interaction between the spread of COVID-19 and the state-level mobility factors and restriction policies. In addition, we show that our approach learns the “bias” linked to state-level demographic characteristics.

### 3.1 State-level Epidemic Forecasting Model

Our approach (Figure 1) trains a model in a pure data-driven manner for each state in the US, including the District of Columbia (DC), to make Δ*t*-ahead prediction of the state-level daily transmission rate 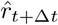 using the state-level daily mobility statistics and government policies at time *t*. The estimated daily confirmed cases in this state can then be derived from the corresponding estimated 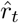 in an accumulated manner. Furthermore, a calibration step is proposed to adjust for short-term changes in population behaviors. We abuse the term “state” a little bit to indicate a state or the DC throughout the paper. The hyper-parameter Δ*t* specifies how far in the future a prediction is made, and is automatically adjusted for each state. The detailed description of the model is provided in Appendix A. The following lists the data used in this work:

**Figure 1:**
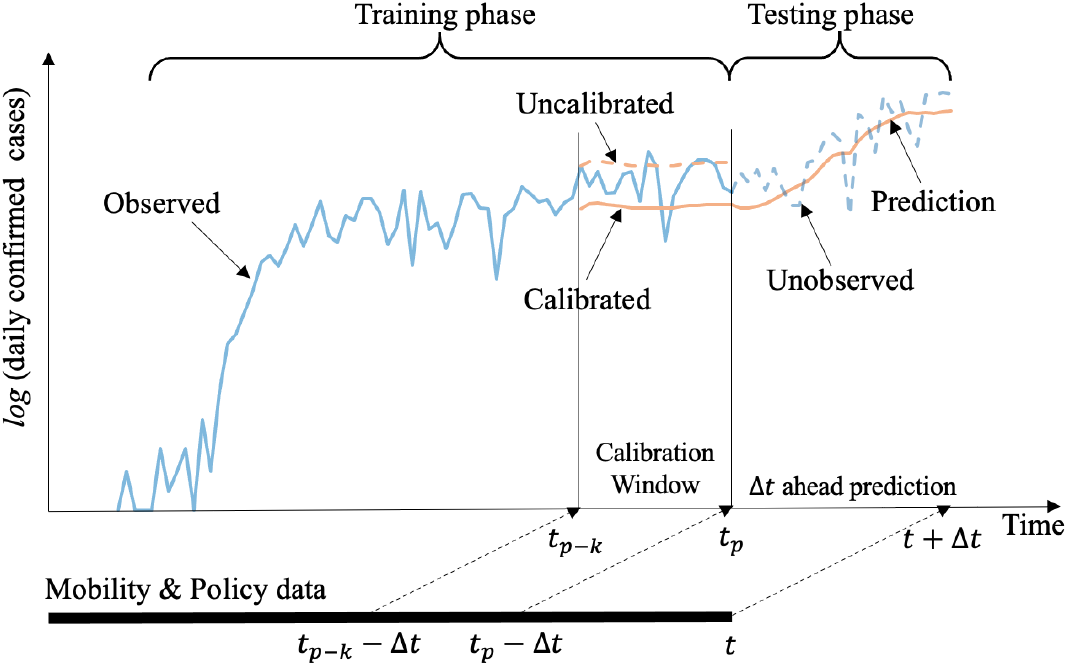
Illustration of the proposed approach for predicting the state-level spread of COVID-19. The *x*-axis represent time. The *y*-axis represents the number of daily cases in logarithmic scale. Our approach makes Δ*t*-ahead predictions, starting at time *t*_*p*_, using the state-level daily mobility statistics and policies. The model calibration is done using the daily confirmed case data within the time window [*t*_*p*−*k*_, *t*_*p*−1_].

- **COVID-19 daily confirmed case data:** The latest COVID-19 daily cases data was obtained from The New York Times^1^.
- **State-level mobility data:** The trip-by-distance mobility data is made available by the Maryland Transportation Institute and Center for Advanced Transportation Technology Laboratory at the University of Maryland^2^. The daily trips are grouped into 10 categories based on the travel distances and another *Staying-at-home* category indicates the ratio of population mostly at home.
- **State restriction policy:** The information about state-level restrictions were extracted from “The Coronavirus Outbreak” forum on the New York Times^3^. This work considers the mask policy and the restaurant restriction policy.
- **State-level demographic information:** This data includes the state-level population density information published by the U.S. Census Bureau^4^, the race structure information (fraction of 7 different race categories) collected by the COVID Tracking Project^5^, and the age structure information (fraction of 6 non-overlapping age groups as well as the high risk population) collected by the Kaiser Family Foundation^6^.

The whole dataset contains the daily confirmed cases (01/21/2020 – 12/08/2020) in the 51 states of the US. The data was split into a training set (01/21/2020 – 11/24/2020) and a test set (11/25/2020 – 12/08/2020). Fifty states issued the restaurant policies, and 34 states issued the public mask policies. For each state, we fit a model using the train data starting from its pandemic start date to 11/24/2020. The pandemic start date of a state is decided in the way discussed in Section A.1. The only hyper-parameter of the model is Δ*t*, which is related to the incubation time of COVID-19 and the efficiency of a state’s healthcare system. The typical incubation period for COVID-19 is around 14 days according to the CDC. Since there were delays in taking tests and reporting cases, we limited Δ*t* to between 15 and 20, and applied ten-fold cross-validation using the training data to determine the optimal Δ*t* of each state. In the test phase, we first smoothed the predicted transmission rate, 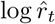, by taking an exponential moving average on the previous 3 days. Then we conduct the calibration step (see Section A.3) using the data between 11/18/2020 and 11/24/2020.

### 3.2 Prediction evaluation metrics

We evaluate the prediction performance based on normalized RMSE (nRMSE):

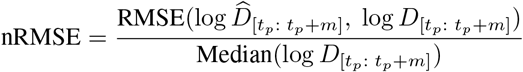

where the prediction period is [*t*_*p*_, *t*_*p*_ +*m*], *m* is set to be no more than 14 in this work, 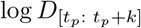 indicates the logarithm of the daily confirmed cases between *t*_*p*_ and *t*_*p*_ +*m*, and 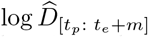 indicates the logarithm of the predicted daily confirmed cases. nRMSE estimates the relative deviation from the the local COVID-19 trend. A value of 0.01 represents the average prediction deviates 1% from the true local trend in a logarithmic scale. We also report the relative accumulated log error (RALE) of cases during [*t*_*p*_, *t*_*p*_ + *m*]:

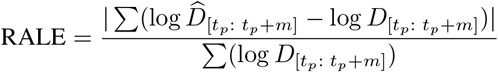

RALE captures the relative deviation from the accumulated cases within *m* days. A value of 0.01 represents the predictive cases deviates 1% from the true cases within *m* days in a logarithmic scale.

#### 3.2.1 Summary of the prediction performance

We ran our approach to make the 3-, 7-, 10- and 14-day predictions in all states. The results are summarized in Figure 2 with details in Table 2. The median nRMSE for 14-day forecasting is 0.035 (i.e., the prediction deviates ≈3.5% from the real local trend at a logarithmic scale). Most nRMSEs of the 14-day forecasting results are within 0.05, indicating that our model works well for most states in the US. The RALE results show a similar trend. We observe that both nRMSE and RALS increase slightly with the forecasting time extends. For instance, the median values of the 3- and 14-day nRMSEs are 0.027 and 0.035, respectively. Figure 3 visualizes the 14-day predictions (both the transmission rates and the daily confirmed cases) of two states, NY (nRMSE = 0.0105, RALE = 0.0036) and LA (nRMSE = 0.0755, RALE = 0.0710). The prediction results on NY and LA are among the best and worst, respectively.

**Table 2:**
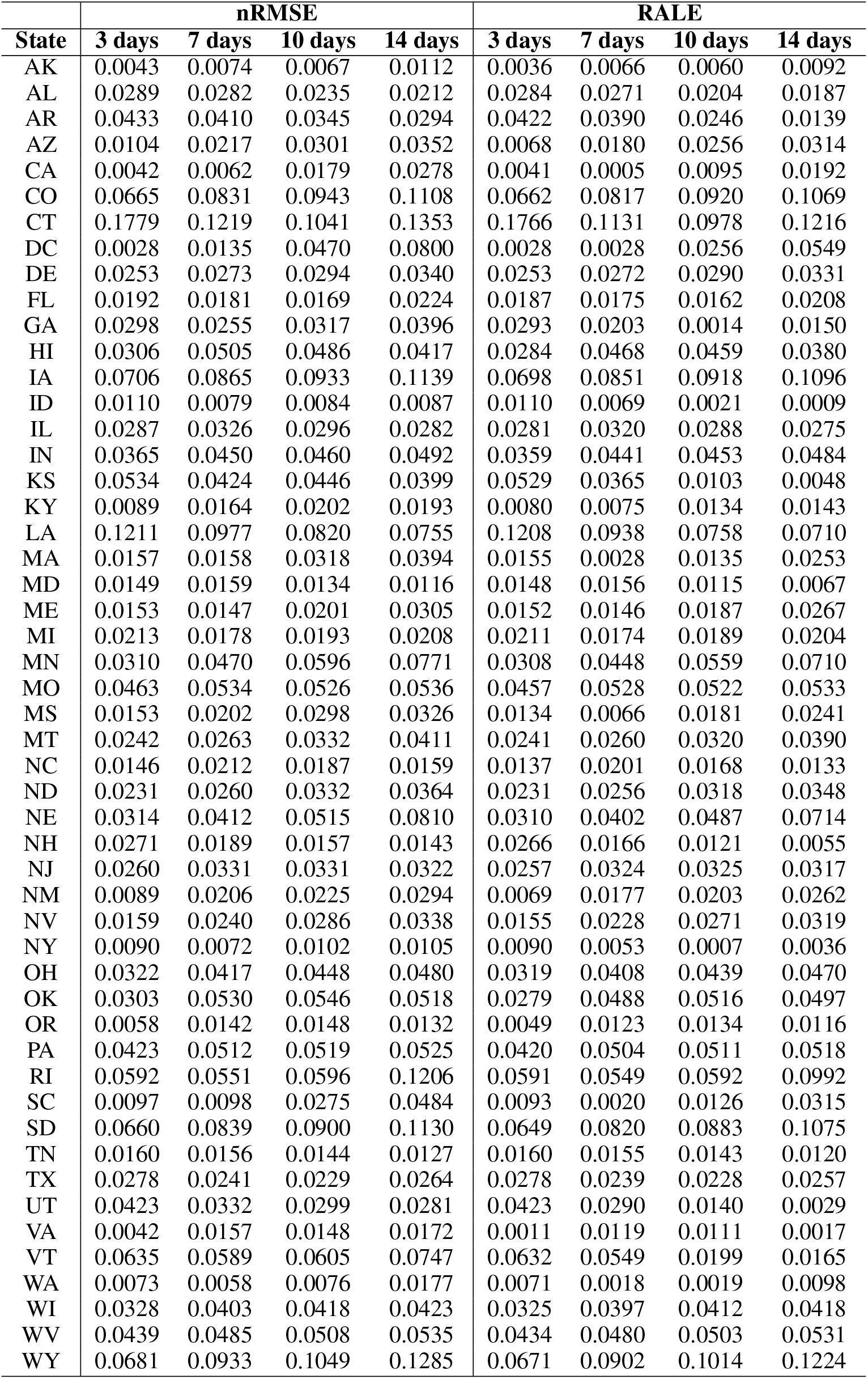
The nRMSE and RALE values of the 3-, 7-, 10-, 14-day state-level predictions.

| State | nRMSE |  |  |  | RALE |  |  |  |
| --- | --- | --- | --- | --- | --- | --- | --- | --- |
|  | 3 days | 7 days | 10 days | 14 days | 3 days | 7 days | 10 days | 14 days |
| AK | 0.0043 | 0.0074 | 0.0067 | 0.0112 | 0.0036 | 0.0066 | 0.0060 | 0.0092 |
| AL | 0.0289 | 0.0282 | 0.0235 | 0.0212 | 0.0284 | 0.0271 | 0.0204 | 0.0187 |
| AR | 0.0433 | 0.0410 | 0.0345 | 0.0294 | 0.0422 | 0.0390 | 0.0246 | 0.0139 |
| AZ | 0.0104 | 0.0217 | 0.0301 | 0.0352 | 0.0068 | 0.0180 | 0.0256 | 0.0314 |
| CA | 0.0042 | 0.0062 | 0.0179 | 0.0278 | 0.0041 | 0.0005 | 0.0095 | 0.0192 |
| CO | 0.0665 | 0.0831 | 0.0943 | 0.1108 | 0.0662 | 0.0817 | 0.0920 | 0.1069 |
| CT | 0.1779 | 0.1219 | 0.1041 | 0.1353 | 0.1766 | 0.1131 | 0.0978 | 0.1216 |
| DC | 0.0028 | 0.0135 | 0.0470 | 0.0800 | 0.0028 | 0.0028 | 0.0256 | 0.0549 |
| DE | 0.0253 | 0.0273 | 0.0294 | 0.0340 | 0.0253 | 0.0272 | 0.0290 | 0.0331 |
| FL | 0.0192 | 0.0181 | 0.0169 | 0.0224 | 0.0187 | 0.0175 | 0.0162 | 0.0208 |
| GA | 0.0298 | 0.0255 | 0.0317 | 0.0396 | 0.0293 | 0.0203 | 0.0014 | 0.0150 |
| HI | 0.0306 | 0.0505 | 0.0486 | 0.0417 | 0.0284 | 0.0468 | 0.0459 | 0.0380 |
| IA | 0.0706 | 0.0865 | 0.0933 | 0.1139 | 0.0698 | 0.0851 | 0.0918 | 0.1096 |
| ID | 0.0110 | 0.0079 | 0.0084 | 0.0087 | 0.0110 | 0.0069 | 0.0021 | 0.0009 |
| IL | 0.0287 | 0.0326 | 0.0296 | 0.0282 | 0.0281 | 0.0320 | 0.0288 | 0.0275 |
| IN | 0.0365 | 0.0450 | 0.0460 | 0.0492 | 0.0359 | 0.0441 | 0.0453 | 0.0484 |
| KS | 0.0534 | 0.0424 | 0.0446 | 0.0399 | 0.0529 | 0.0365 | 0.0103 | 0.0048 |
| KY | 0.0089 | 0.0164 | 0.0202 | 0.0193 | 0.0080 | 0.0075 | 0.0134 | 0.0143 |
| LA | 0.1211 | 0.0977 | 0.0820 | 0.0755 | 0.1208 | 0.0938 | 0.0758 | 0.0710 |
| MA | 0.0157 | 0.0158 | 0.0318 | 0.0394 | 0.0155 | 0.0028 | 0.0135 | 0.0253 |
| MD | 0.0149 | 0.0159 | 0.0134 | 0.0116 | 0.0148 | 0.0156 | 0.0115 | 0.0067 |
| ME | 0.0153 | 0.0147 | 0.0201 | 0.0305 | 0.0152 | 0.0146 | 0.0187 | 0.0267 |
| MI | 0.0213 | 0.0178 | 0.0193 | 0.0208 | 0.0211 | 0.0174 | 0.0189 | 0.0204 |
| MN | 0.0310 | 0.0470 | 0.0596 | 0.0771 | 0.0308 | 0.0448 | 0.0559 | 0.0710 |
| MO | 0.0463 | 0.0534 | 0.0526 | 0.0536 | 0.0457 | 0.0528 | 0.0522 | 0.0533 |
| MS | 0.0153 | 0.0202 | 0.0298 | 0.0326 | 0.0134 | 0.0066 | 0.0181 | 0.0241 |
| MT | 0.0242 | 0.0263 | 0.0332 | 0.0411 | 0.0241 | 0.0260 | 0.0320 | 0.0390 |
| NC | 0.0146 | 0.0212 | 0.0187 | 0.0159 | 0.0137 | 0.0201 | 0.0168 | 0.0133 |
| ND | 0.0231 | 0.0260 | 0.0332 | 0.0364 | 0.0231 | 0.0256 | 0.0318 | 0.0348 |
| NE | 0.0314 | 0.0412 | 0.0515 | 0.0810 | 0.0310 | 0.0402 | 0.0487 | 0.0714 |
| NH | 0.0271 | 0.0189 | 0.0157 | 0.0143 | 0.0266 | 0.0166 | 0.0121 | 0.0055 |
| NJ | 0.0260 | 0.0331 | 0.0331 | 0.0322 | 0.0257 | 0.0324 | 0.0325 | 0.0317 |
| NM | 0.0089 | 0.0206 | 0.0225 | 0.0294 | 0.0069 | 0.0177 | 0.0203 | 0.0262 |
| NV | 0.0159 | 0.0240 | 0.0286 | 0.0338 | 0.0155 | 0.0228 | 0.0271 | 0.0319 |
| NY | 0.0090 | 0.0072 | 0.0102 | 0.0105 | 0.0090 | 0.0053 | 0.0007 | 0.0036 |
| OH | 0.0322 | 0.0417 | 0.0448 | 0.0480 | 0.0319 | 0.0408 | 0.0439 | 0.0470 |
| OK | 0.0303 | 0.0530 | 0.0546 | 0.0518 | 0.0279 | 0.0488 | 0.0516 | 0.0497 |
| OR | 0.0058 | 0.0142 | 0.0148 | 0.0132 | 0.0049 | 0.0123 | 0.0134 | 0.0116 |
| PA | 0.0423 | 0.0512 | 0.0519 | 0.0525 | 0.0420 | 0.0504 | 0.0511 | 0.0518 |
| RI | 0.0592 | 0.0551 | 0.0596 | 0.1206 | 0.0591 | 0.0549 | 0.0592 | 0.0992 |
| SC | 0.0097 | 0.0098 | 0.0275 | 0.0484 | 0.0093 | 0.0020 | 0.0126 | 0.0315 |
| SD | 0.0660 | 0.0839 | 0.0900 | 0.1130 | 0.0649 | 0.0820 | 0.0883 | 0.1075 |
| TN | 0.0160 | 0.0156 | 0.0144 | 0.0127 | 0.0160 | 0.0155 | 0.0143 | 0.0120 |
| TX | 0.0278 | 0.0241 | 0.0229 | 0.0264 | 0.0278 | 0.0239 | 0.0228 | 0.0257 |
| UT | 0.0423 | 0.0332 | 0.0299 | 0.0281 | 0.0423 | 0.0290 | 0.0140 | 0.0029 |
| VA | 0.0042 | 0.0157 | 0.0148 | 0.0172 | 0.0011 | 0.0119 | 0.0111 | 0.0017 |
| VT | 0.0635 | 0.0589 | 0.0605 | 0.0747 | 0.0632 | 0.0549 | 0.0199 | 0.0165 |
| WA | 0.0073 | 0.0058 | 0.0076 | 0.0177 | 0.0071 | 0.0018 | 0.0019 | 0.0098 |
| WI | 0.0328 | 0.0403 | 0.0418 | 0.0423 | 0.0325 | 0.0397 | 0.0412 | 0.0418 |
| WV | 0.0439 | 0.0485 | 0.0508 | 0.0535 | 0.0434 | 0.0480 | 0.0503 | 0.0531 |
| WY | 0.0681 | 0.0933 | 0.1049 | 0.1285 | 0.0671 | 0.0902 | 0.1014 | 0.1224 |

**Figure 2:**
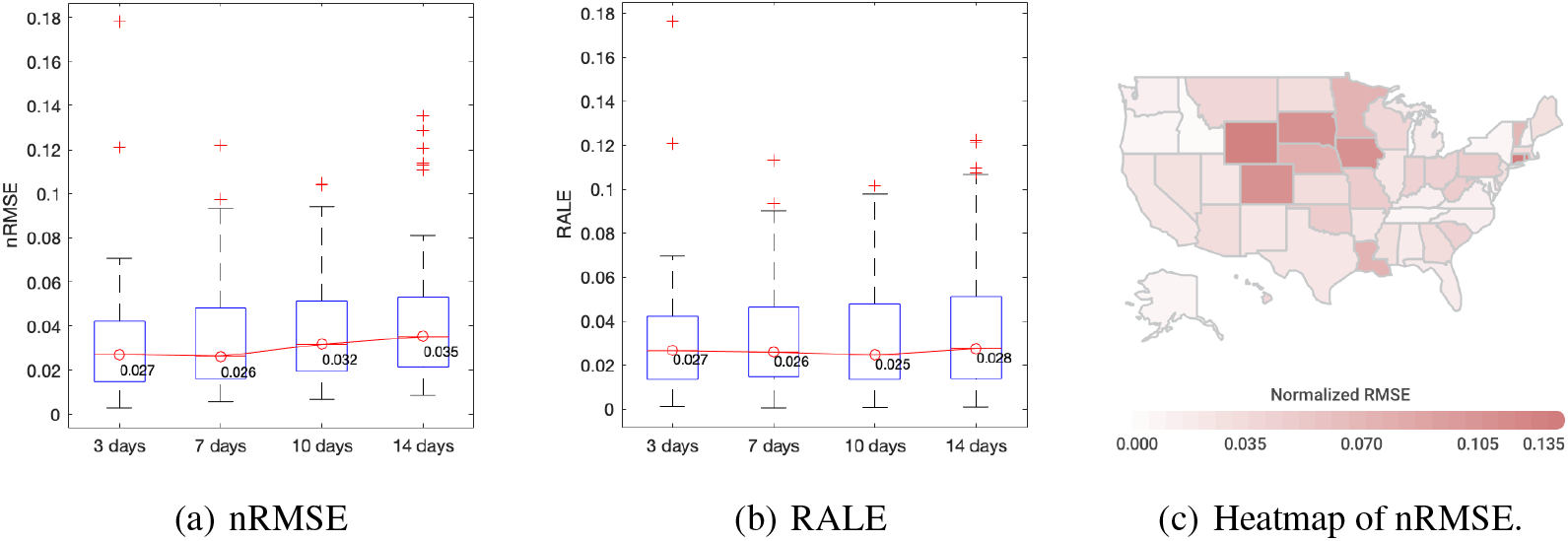
The summary of the 3-, 7-, 10- and 14-day state-level forecasting performance. (a) The state-level nRMSE values. (b) The state-level RALE values. (c) The geographic heatmap of the 14-day forecasting nRMSE.

**Figure 3:**
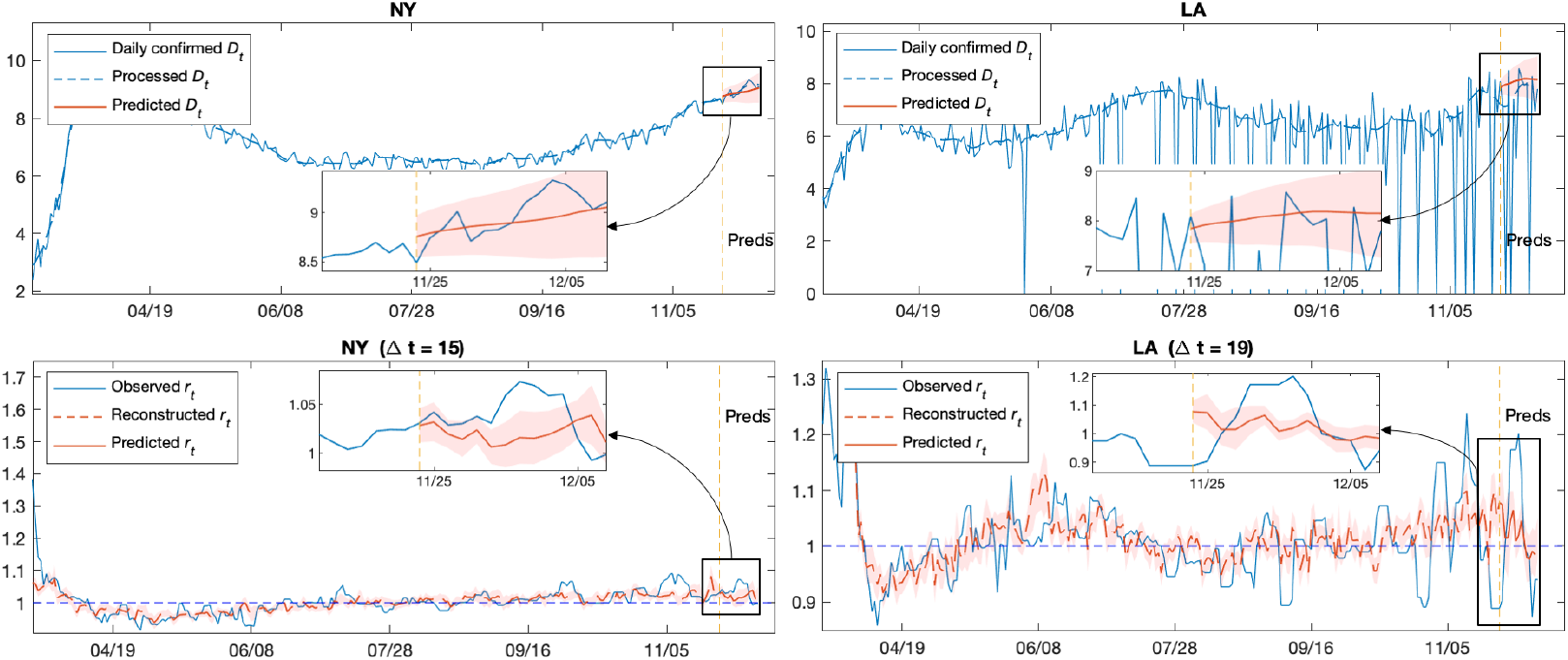
The 14-day predictions of the COVID-19 transmission rates and daily cases in NY (nRMSE = 0.0105, RALE = 0.0036) and LA (nRMSE = 0.0755, RALE = 0.0710) in logarithmic scale. All *x*-axis indicate time. The *y*-axes in the top plots indicate the logarithm of the daily confirmed cases. The *y*-axes in the bottom plots indicate the transmission rate values. The yellow dash vertical lines indicate the starting points of the prediction periods. The blowouts highlight the predictions. The red shaded areas indicate the 95% confidence intervals.

We should point out that a large nRMSE or RALE may indicate large volatility in the confirmed daily cases due to delay in reporting, rather than the weak performance of the corresponding model. Figure 10 shows four representative states that contain significant volatilities in daily confirmed cases in or after the forecasting period (11/25/2020 – 12/08/2020). However, their overall future trends match our forecasting curves very well.

### 3.3 Significant factors in predicting COVID-19 trend

The predictors have different levels of effects on COVID-19 trend prediction across the US (see Figure 4b-f, with more details in Figure 8). Using 0.05 as the *p*-value cutoff, we observed several factors are frequently identified as significant across the states (Figure 4a). *Mask Policy* has the highest frequency of 0.7647 (26 out of 34 states that issued the mask policy), indicating that the mask policy has the most impact on the change of epidemic dynamics. The estimated coefficients of *Mask Policy* and three other significant factors (*Restaurant Policy, Stay-at-home* and *Dis-0-1*) are negative in nearly all cases (Appendix B), which indicates these factors help hamper the spread of COVID-19. This is consistent with how people and governments fight against COVID-19: wearing masks, closing restaurants and staying at home. Mobility categories *Dis-1-3* and *Dis*>*500* also have general impacts. However, their estimated coefficients are positive in most cases, indicating that they help promote the spread of COVID-19. We suspect that *Dis-1-3* may correspond to walking within local communities and that *Dis*>*500* mostly represents cross-state travels.

**Figure 4:**
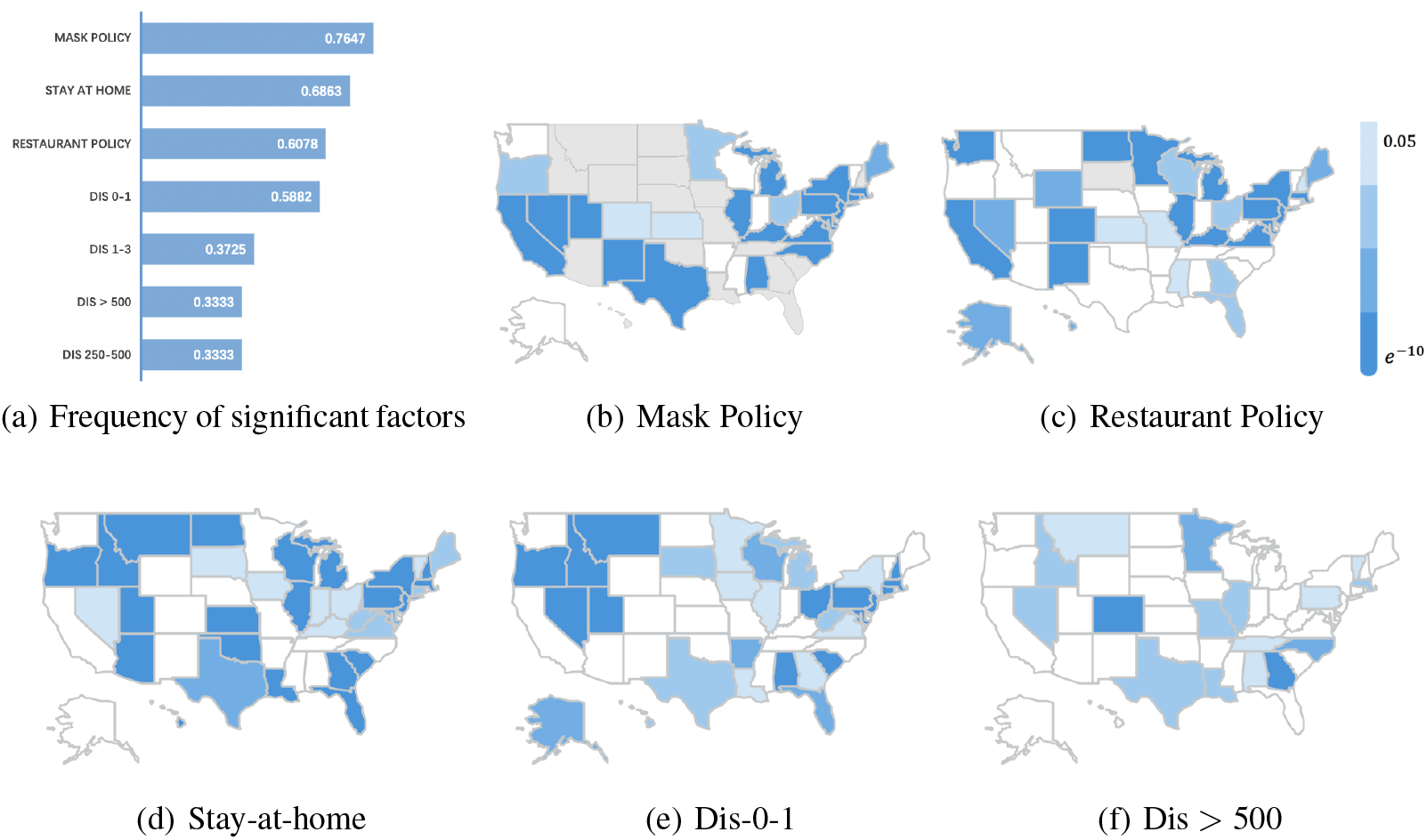
(a) Frequency of each factor identified to be significant within states. Factor with high frequency implies its general influences on most states. (b)–(f) Heatmaps of each factor’s p-values. Here a state is colored grey if it doesn’t incorporate such factor into regression. Remaining results are also reported in Figure 8 in the Appendix.

Interestingly, other mobility categories become significant for a few states. For example, the long distance mobility categories (*Dis-250-500* and *Dis*>*500*) are both significant in regression for states NV and MT. This might be explained by the low population densities in those states. There may be other very complex interactions between mobility and demographic properties, which we leave for future exploration.

### 3.4 Demographic interpretation of the state-level biases in COVID-19 transmission

We trained one daily case prediction model for each state (including the DC) and obtained 51 models in total. Notice that the intercept term in each model represents the baseline transmission rate of the corresponding state. We hypothesized that the differences in the baseline transmission rates were due to the state-level demographics (e.g., population density, age structure, race structure, etc.).

To investigate this, we used lasso (Tibshirani, 1996) to select four demographic variables highly related to the state-level model biases, which include *Top Density*^7^ (*p*-value = 0.0293), *Adults-35-54* (*p*-value = 0.1003), *Hispanic-Or-Latino* (*p*-value = 0.0112) and *AmericanIndian-Or-AlaskaNative* (*p*-value = 0.1507). We then used them to reconstruct the baseline transmission rates using nonregularized linear regression (see Table 1 and Figure 5). Our findings resonate with the findings in previous works that the spread of COVID-19 were highly relevant to population densities (Rocklöv & Sjödin, 2020) and ethnic minorities (Dyer, 2020; Kirby, 2020), and hence aid in understanding the spread of COVID-19 and increase the interpretability of our model.

**Table 1:** Regression Table on baseline transmission rates

|  | Estimate | SE | tStat | pValue |
| --- | --- | --- | --- | --- |
| (Intercept)* | 0.63678 | 0.20508 | 3.1051 | 0.0032527 |
| Top-density | 0.013506 | 0.006005 | 2.2492 | 0.029331 |
| Adults-35-54 | 1.4687 | 0.8757 | 1.6771 | 0.1003 |
| Hispanic-Or-Latino | 0.21254 | 0.080443 | 2.6421 | 0.011223 |
| AmericanIndian-Or-AlaskaNative | -0.21076 | 0.14421 | -1.4614 | 0.1507 |

\* This is the intercept of regression on demographics.
Number of observations: 51, Error degrees of freedom: 46
Root Mean Squared Error: 0.0519
R-squared: 0.421, Adjusted R-Squared: 0.391
F-statistic vs. constant model: 5.34, p-value = 1.83e-05

**Figure 5:**
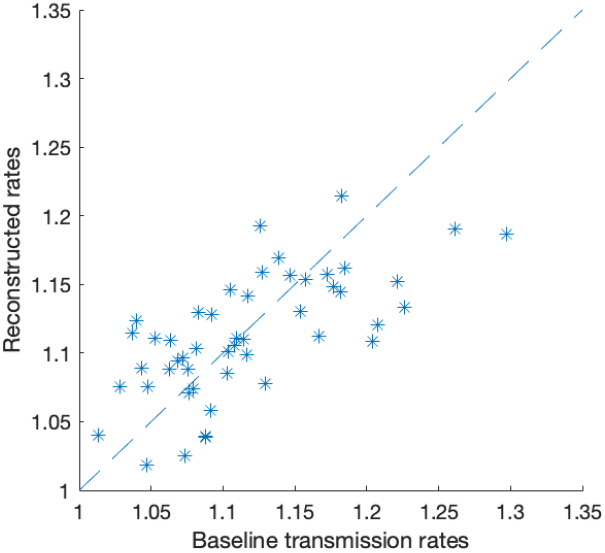
Daily transmission rate is estimated by time-dependent mobility variables as well as categorical variables for policies. Then daily cases are predicted from previous transmission rates. The biased term (intercept) from the regression model is the baseline transmission rate under normal mobility and conditions before the pandemic. The baseline rate is highly related to each state’s demographics.

## 4 Conclusions

In this paper, we propose a data-driven approach that trains regression models for forecasting the state-level COVID-19 daily transmission rates using the state-level mobility data and restrictive policies. The transmission rates can then be used to estimate the daily confirmed cases in an accumulated manner. Our approach uses a calibration step to adjust for short-term changes in population behaviors. Our empirical study results show that the proposed approach can reliably and accurately forecast (2 weeks ahead) the state-level COVID-19 spread. We also studied statistically significant factors as well as their impacts on the COVID-19 pandemic, and the findings allow us to better understand how population mobility and government policies may affect the spread of COVID-19. Our prediction results can be used by local governments or healthcare systems to prepare ahead, and the discovered quantitative relationships between COVID-19 and population mobility as well as policies can be used to by policymakers in devising customized response strategies.

## Data Availability

All data are available in this manuscript.

https://github.com/nytimes/covid-19-data

https://data.bts.gov/Research-and-Statistics/Trips-by-Distance/w96p-f2qv

https://github.com/yifeiwang15/COVID-19-Mobility/tree/main/data

## Code and Data Availability

Our codes and the data used in this work are available on GitHub at https://github.com/yifeiwang15/COVID-19-Mobility.

## Author Contributions

Yifei Wang developed the whole model architecture, planned and carried out the experiments, and mainly wrote the manuscript. Pengyu Hong initialized this project, conceived of the presented idea, supervised the experiments and revised the manuscript. Michael Peng did data extraction as well as data filtering and helped improve the manuscript. Michael Peng and Frank Liu developed a website (https://broad-well.github.io/covid-trend-prediction/) to visualize our predictions on all states. Long Sha participated in brainstorms and discussions.

## Acknowledgments

This work was partially supported by NSF OAC 1920147.

## A Methods

### A.1 Data and preprocessing

#### COVID-19 daily case data

The COVID-19 data used in this work is the US state-level daily confirmed cases, denoted as *D*_*t*_ where *t* is date. The data is very noisy, especially in the beginning of the pandemic, due to various reasons, such as, delay in reporting, and so on. We performed the following preprocessing. For each state, we first detected the pandemic start time *t*_*s*_ as the first day of the first three consecutive days with non-zero daily confirmed cases. We then smoothed *D*_*t*_ after *t*_*s*_ by taking a moving median using a sliding window of size 7 followed by a moving average using a sliding window of size 5. In the rest of the paper, *D*_*t*_ refers to the preprocessed daily cases.

#### Mobility data

The state-level travel statistics are daily aggregates of residents’ movements based on their mobile phone data, and provide information about population mobility. A trip was counted if a person stayed away from home for more than 10 minutes. The daily trips were grouped into 11 categories based on their travel distances (see Figure 6 for an example). For instance, the *Dis-5-10* category indicates the number of trips within the range of 5-10 miles. The *Staying-at-home* category records the size of the population that did not stay away from home for more than 10 minutes. In each state, the *Staying-at-home* category data was normalized by the state population, and other categories were normalized by the state population not staying at home. Each category is then standardized to represent the relative changes in mobility from the pre-pandemic level. This was done by subtracting the median pre-pandemic mobility value and then dividing the maximal pre-pandemic mobility value.

**Figure 6:**
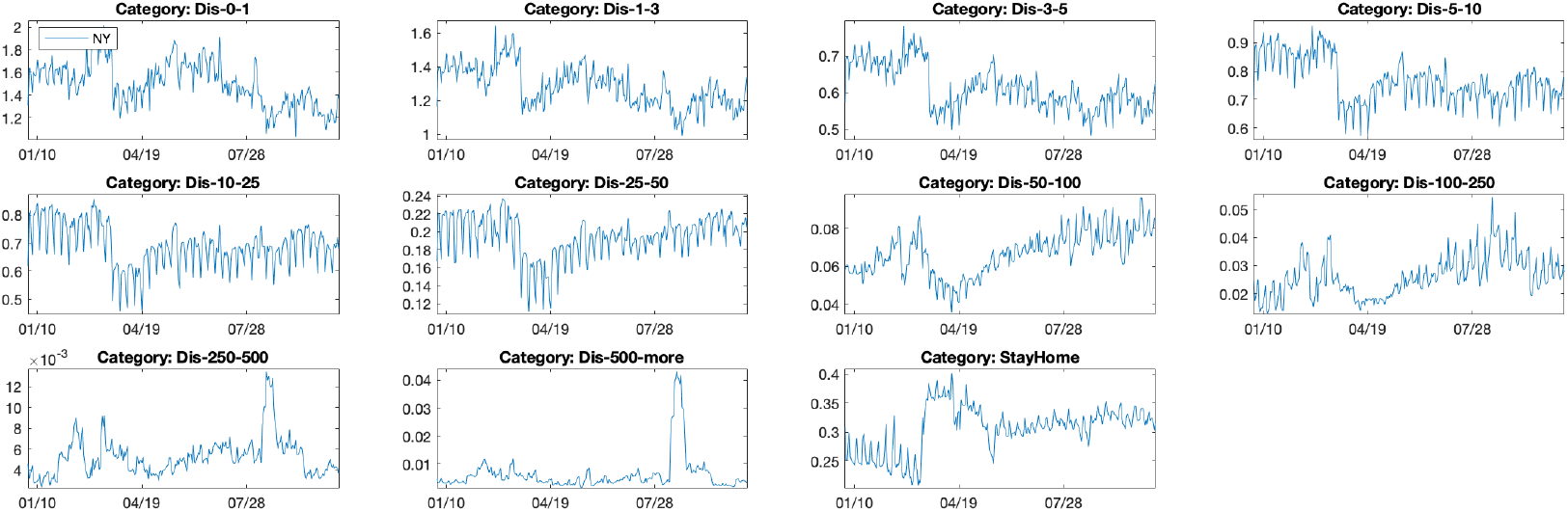
The normalized mobility data of the New York state. There are 11 categories based on the travel distances. The normalization procedure is explained in Section A.1.

There exists abnormal activities across the US around the later summer 2020 when schools start and in early November 2020 when the election was held. These sudden irregular travel patterns were associated with distinct yet unknown population behaviors. We suspect that the subpopulation, which exerted the abnormal travel patterns, deployed special the required protection/quarantine means and hence contributed little to the spread of COVID-19. Hence, the corresponding mobility data should not be used without appropriate processing in training the models and making predictions. To this end, we detected outliers as samples more than three scaled median absolute deviations away from the median. Then, we conducted Principal Component Analysis on the training data and reconstructed the detected outliers with the first 4 principle components. Figure 7 shows an example of outlier detection and reconstruction in the mobility data category *Dis* > *500* of the New York and California states.

**Figure 7:**
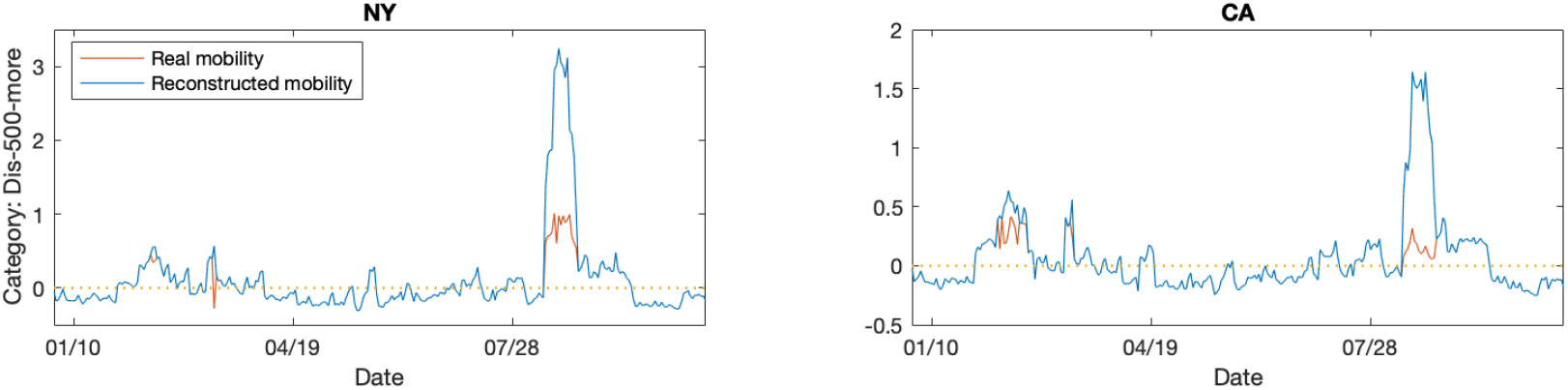
There is a national spike of long-distance travels (the *Dis*>*500* mobility category) in mid-August, which might be associated with the starts of schools/universities. The data of two states (NY and CA) are shown as examples. The abnormal mobility samples are “corrected” using Principal Component Analysis as described in Section A.1.

#### State restriction policy

We considered the state-level mask wearing policy and restaurant opening policy as two binary variables. If a policy is instated, the value of its variable is 1, otherwise 0.

#### Demographic information

The following state-level demographic information, which was also used in Yang et al. (2020a): (i) local population density, (ii) local age structure (non-overlapping age groups), (iii) local race structure (different race categories).

### A.2 Regression model for epidemic prediction

The epidemic transmission rate in each state at time *t* is defined as the ratio between the state-level confirmed daily cases at *t* and that at *t*−1, i.e., *r*_*t*_ = *D*_*t*_*/D*_*t*− 1_, where *D*_*t*_ is the number of the daily confirmed cases at time *t*. The transmission rate *r*_*t*_ can be algebraically mapped to the reproduction number in epidemiology (Fan et al., 2020). Assuming no auto-correlation in transmission rates, we first use a robust linear regression technique (Holland & Welsch, 1977) to estimate the logarithm of the state-level transmission rate (i.e., 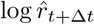) using the mobility statistics and policies at time *t*, where the hyper-parameter Δ*t* specifies how far in the future a prediction is made and its optimal value can be adjusted using ten-fold cross-validation. Assuming prediction starts at time *t*_*p*_, the logarithm of the predicted daily cases 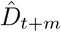, where *m* ≤ Δ*t*, can be derived using the estimated transmission rates as follows:

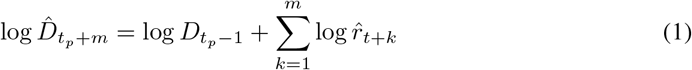

where 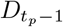 is the ground-truth number of the daily cases at time *t*_*p*_ − 1.

### A.3 Calibrating the forecast

The above regression model assumes stationary relationships between the transmission rate variable and the predictors (i.e., population mobility and policies), which is not necessarily true in reality. For example, it is well known that population behaviors (e.g., awareness of conditions relating to public health, compliance to policies, etc.) vary over time, which can contribute to changes in transmission rates. Moreover, the reporting error associated with the daily confirmed cases (i.e., 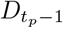 in eq. 1) and the accumulated prediction error in 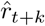 can degenerate the predictions of the daily cases.

Hence, we introduce a calibration step that uses the data in a short window immediately preceding the forecast window (see Figure 1) to make adjustments. This step makes a reasonable assumption that the relationships between the transmission rate variable and its predictors remain unchanged over a short time period composed of the calibration and forecast windows. Assume prediction should start at time *t*_*p*_, we use the time window [*t*_*p*−*k*_, *t*_*p*−1_] to linearly calibrate the model trained by using the data up to *t*_*p*−1_ as

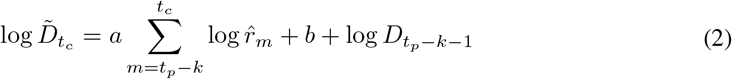

where 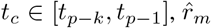 is the output of the pre-calibrated model, and *a* & *b* are two calibration parameters. The hyper-parameter *k* controls the attention span of the calibration step. A small *k* direct the calibration step to focus on short-term epidemic trends, and vice versa. The parameter *a* accounts for the time-changing relationship between the transmission rate and its predictors, and *b* accounts for both the uncertainty associated with 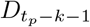 and the error in forecasting *r*_*t*_. These two parameters can be solved by optimizing

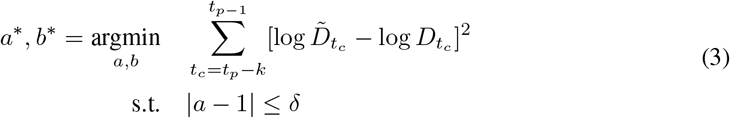

where *δ* > 0 controls the maximal scaling effect to prevent numerical instability in estimating *a*^*∗*^ due to the large uncertainties associated with reports of daily cases. We set *δ* = 0.01 in our experiments. After calibration, the prediction at *t* ≥ *t*_*p*_ should be calculated as 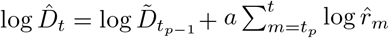, where 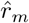 is the Δ*t*-step-ahead prediction made by the pre-calibrated model.

## B Additional results

**Figure 8:**
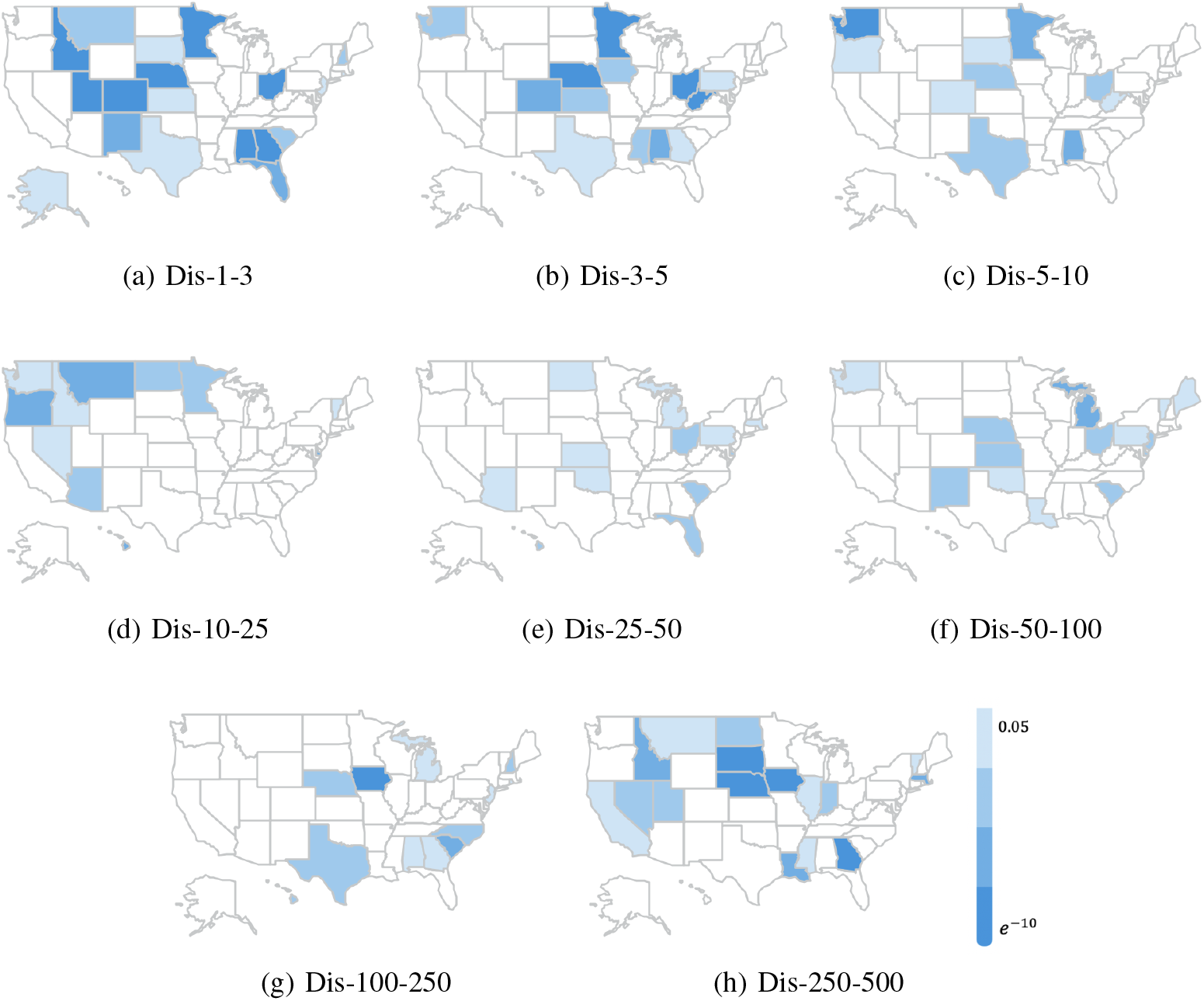
The regression *p*-value heatmaps of the mobility data categories.

**Figure 9:**
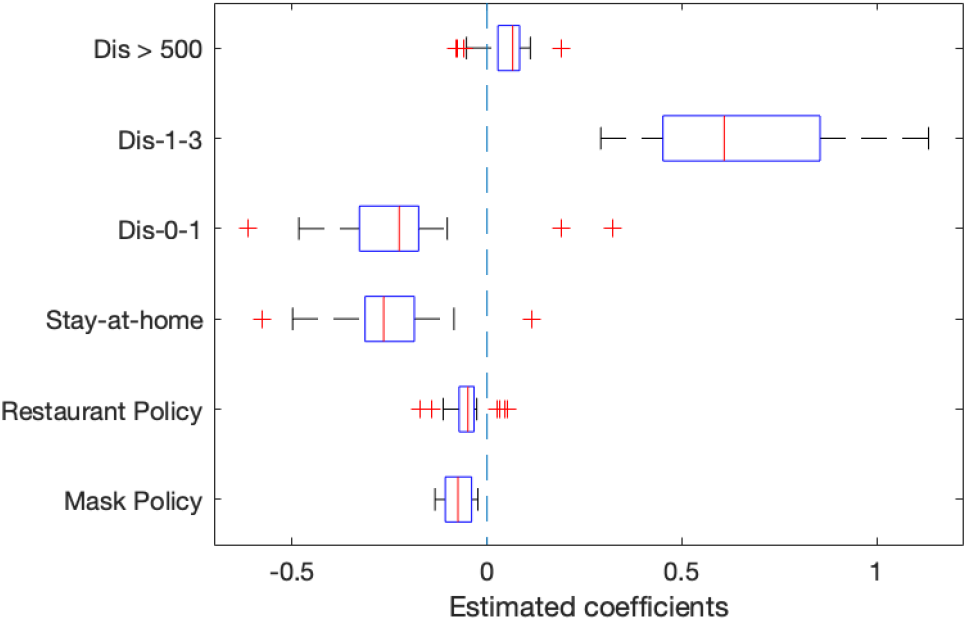
The most generic significant factors identified by our approach (see Figure 4(a)) as well as their descriptive statistics of estimated coefficients, according to 51 state-level regression models. Only coefficients with significant level of 0.05 are included in the box plot. The coefficients of *Mask Policy, Restaurant Policy, Stay-at-home* and *Dis-0-1* almost take negative values, showing stable negative correlations with the transmissions rates and indicating that they help prevent the spread of COVID-19. In contrast, the coefficients of *Dis-1-3* and *Dis*>*500* almost take positive values, showing stable positive correlations with the transmissions rates and indicating that frequent short-distance travels and cross-state travels help promote the spread.

**Figure 10:**
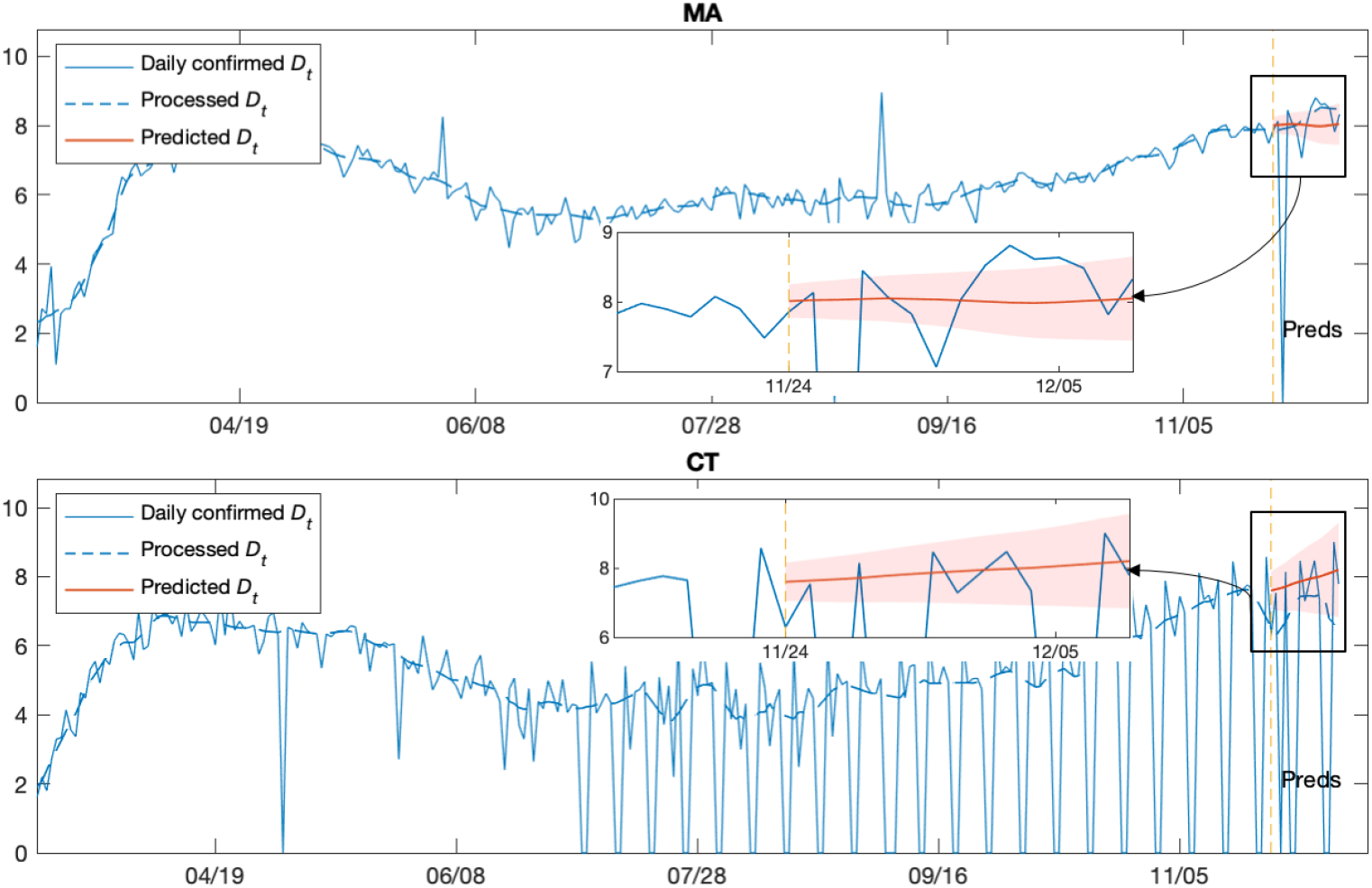
Two representative cases show that the trained models can forecast the trends very well even though they have relatively large prediction nRMSE and RALE: MA (nRMSE = 0.0394, RALE = 0.0253) and CT (nRMSE = 0.1353, RALE = 0.1216). These large nRMSE/RALE values are due to the delays or skips in reporting the COVID-19 daily confirmed cases by the corresponding states. These results indeed indicate the robustness and reliability of our approach. The *x*-axis indicate time. The *y*-axes indicate the logarithm of the daily confirmed cases. The yellow dash vertical lines indicate the starts of the prediction periods. The blowouts highlight the predictions. The red shaded areas indicate the 95% confidence intervals.

## Footnotes

1 https://github.com/nytimes/covid-19-data

2 https://data.bts.gov/Research-and-Statistics/Trips-by-Distance/w96p-f2qv

3 https://www.nytimes.com/interactive/2020/us/states-reopen-map-coronavirus.html

4 https://www.census.gov

5 https://covidtracking.com

6 https://www.kff.org/other/state-indicator/distribution-by-age/?currentTimeframe=0&sortModel=%7B%22colId%22:%22Location%22,%22sort%22:%22asc%22%7D%23notes

7 The highest population density within state.

